# What is the efficacy and safety of rapid exercise tests for exertional desaturation in Covid-19: A rapid review protocol

**DOI:** 10.1101/2020.10.31.20223453

**Authors:** Asli Kalin, Babak Javid, Matthew Knight, Matt Inada-Kim, Trisha Greenhalgh

## Abstract

**Background:** Even when resting pulse oximetry is normal in the patient with acute Covid-19, hypoxia can manifest on exertion. We sought to summarise the literature on the performance of different rapid tests for exertional desaturation.

**Research question:** What tests have been formally evaluated for the rapid assessment of exertional hypoxia? What is the evidence for their accuracy, practicability and safety in the context of suspected acute Covid-19? To what extent will these tests help identify patients with evidence of either silent or hidden hypoxia leading to earlier recognition of those at risk of severe outcomes?

**Method:** We aim to review three independent searches of AMED, CINAHL, EMBASE MEDLINE, Cochrane and PubMed using LitCovid, Scholar and Google databases until 24^th^ September 2020. Screening, data abstraction, and quality appraisal of full-text papers will be completed independently by two reviewers including a topic expert and a review expert. Studies will be tabulated and assessed for risk of bias using QUADAS 2 tool.

**Discussion:** This rapid review aims to produce key findings relevant to the assessment of exertional desaturation in patients with suspected Covid-19. Establishing a validated tool to assess exertional desaturation will help to ensure that future research on this topic can be undertaken in a consistent way. An exertional desaturation test should be used in combination with a comprehensive clinical evaluation and only on patients whose resting oximetry reading is 96% or above unless in a supervised care setting. It should be terminated if the patient experiences adverse effects.

## Background

A substantial proportion of patients with acute coronavirus 19 (Covid-19) develop a potentially critical form of the illness requiring intensive care unit admission [1]. The degree of lung involvement in acute Covid-19 is variable, producing a spectrum of illness from mild upper respiratory tract symptoms to acute respiratory distress syndrome [2]. Patients with mild initial symptoms can rapidly deteriorate to severe or critical cases. In hospitalised patients, hypoxaemia and the need for oxygen are independent predictors of severe outcomes [3, 4]. The usual time from symptom onset to the development of severe hypoxemia is between 7 and 12 days [5, 6]. Recent prognostic tools such as the 4C score have emphasised the importance of identifying hypoxia early [3, 7] and there are physiological reasons for managing this hypoxia actively [8, 9].

The poor correlation between both subjective feeling of shortness of breath (dyspnoea) and objective measures of breathlessness and hypoxia in patients with Covid-19 has resulted in UK guidelines recommending that the assessment of the breathless, unwell or high-risk patient should include oximetry [5]. For example, a retrospective cohort study of 64 Covid-19 patients considered eligible for home oximetry monitoring showed that the presence of dyspnoea had a positive predictive value of only 42% for hypoxemia and absence of dyspnoea had a negative predictive value of 86% for excluding it [10].

Indeed, the mismatch between relatively mild subjective respiratory distress and objective evidence of peripheral hypoxia is now recognised as a feature of Covid-19 and has been termed “silent” or “happy” hypoxemia [11, 12]. This mismatch has been attributed to moderate to severe ventilation-perfusion mismatch [11]. Whereas many common lung diseases (such as asthma and chronic obstructive pulmonary disease) produce mainly ventilation defects where air does not reach the alveoli, some (such as interstitial lung disease, sarcoidosis and pulmonary embolism) produce defects of perfusion in which gas transfer across the alveolar-capillary barrier is impaired. Several mechanisms for the perfusion defect in Covid-19, including intra-pulmonary shunting, loss of lung perfusion regulation, intravascular microthrombi or reduced lung compliance, have been proposed [12, 13].

Hypoxia is common in acute severe Covid-19. Richardson et al found that 28% of 5700 patients admitted to hospital with acute Covid-19 in New York City were sufficiently hypoxemic to need supplemental oxygen on admission [14]. Yet dyspnoea was reported in only 18.7% of 1099 hospitalized Covid-19 patients, despite low PaO2/FiO2 ratios and requirement for supplemental oxygen in 41% [15]. In a study of the 19 worst-affected countries worldwide, Goyal et al reported that rates of mortality were significantly higher in those countries where policies recommended lower oxygen saturations before administering supplemental oxygen than in those that aimed to normalised saturations, though there may have been other explanations for these differences including different degrees of preparedness for a pandemic and different incidence rates [16].

It is widely reported that some patients with acute Covid-19 have normal pulse oximetry at rest but their readings deteriorate on exertion (unpublished data). UK national guidance, for example, recommends the use of exertional desaturation tests to detect early deterioration of patients with COVID-19 in community and emergency settings [17-19]. In consensus exercises, front-line clinicians have identified the late transfer of patients with exertional desaturation (i.e. a fall of 3% or more in pulse oximetry reading on exercise) as a possibly remediable cause of poor outcome [20]. In other words, if we could better identify those with exertional desaturation and escalate their care more promptly, we could potentially reduce mortality. The question then becomes: which test should we use to confirm or exclude exertional desaturation?

Several publications recommend or suggest the use of exertional tests in the assessment of COVID-19 patients. Mantha et al discuss the use of a modified 6-minute walk test (6MWT), wearing masks and only in those under 70, on day 4 or 5 of clinical illness to risk-stratify patients [6]. This suggestion is echoed by Noormohammadpour and Abolhasani, who propose that deterioration in the 6MWT test can be used to identify those who need referral to hospital [21]. Pandit et al recommend the 6MWT for those under 60 who are not short of breath at rest and a 3-minute walk test (3MWT) for those over 60 who are unable to complete the longer test, but warn that the test is contra-indicated in patients who are hypoxic at rest (SpO2 < 94%), short of breath at rest, not able to walk unassisted, have Eisenmenger’s syndrome, severe anaemia, unstable angina or valvular heart disease [2]. They suggest that the 6MWT or 3MWT test can be performed at home or in hospital under the supervision of either a family member or a healthcare professional. These authors define exertional hypoxia as an absolute drop in oximeter reading by 3% or more from baseline, and recommend escalation of care for all such patients.

In sum, consensus guidance and editorials recommend tests for exertional hypoxia in Covid-19, but the evidence base for these tests has not previously been formally reviewed. Indeed, the prognostic utility of exertional desaturation remains unknown. Another unknown is the safety of such tests in patients with suspected Covid-19, especially when used remotely without a clinician physically present. Establishing a validated tool to assess exertional desaturation will help to ensure that future research on this topic can be undertaken in a consistent way.

### Review objective and research questions

The overall objective of this rapid review is to examine the published evidence base for the use of rapid exercise tests to confirm or exclude exertional hypoxia in patients with Covid-19. We are particularly interested in tests that could be used outside of the clinical setting, since the reality of acute Covid-19 often involves a remote assessment (with the patient at home at a distance from the clinician) or one in a bespoke ambulatory setting such as a “hot hub” (where exercise tests may be performed outside in car parks, for example, for infection control reasons).

The research questions are:

1. What exercise tests have been used to assess exertional hypoxia at home or in an ambulatory setting in the context of Covid-19 and to what extent have they been validated?
2. What exercise tests have been used to assess exertional hypoxia in other lung conditions and to what extent have they been validated?

## Methods

We are going to follow Cochrane Rapid Review guidelines [22]. Our team includs experienced health librarians as well as systematic reviewers and clinicians (including a general practitioner, general physician, respiratory consultant and emergency medicine consultant with experience of undertaking exercise tests). References will be downloaded to Endnote (version 9.0) to maintain and manage citations and facilitate the review process.

### Search strategy and selection criteria

Three independent searches will be conducted. The first will search LitCovid, Scholar and Google using the terms ‘step test or field test’ and ‘hypoxia or exertional desaturation’. From these results, promising articles will then be used to find additional documents via two methods – forward and backward citation matching, and searching for related articles (also using Microsoft Academic Search).

The second search will covered the following databases: AMED, CINAHL, EMBASE MEDLINE, and PubMed using the over-arching question ‘Very short exercise desaturation tests for use in the emergency department’. Search terms are:

➢ (quick OR short) AND oxygen) AND exercise) AND (desaturation OR saturation)) AND test*(“emergency department*” AND oxygen) AND exercise) AND desaturation) AND test*) (“emergency department*” AND oxygen) AND exercise) AND saturation) AND test*)
➢ (“emergency department*” AND oxygen) AND exercise) AND desaturation)
➢ (“emergency department*” AND oxygen) AND exercise) AND saturation)
➢ (1-min sit-to-stand test) or (“1 min sit to stand test”) or (“one-minute sit to stand”)

The third search will involve searching the Cochrane Database of Systematic Reviews (CDSR), the Cochrane Central Register of Controlled Trials (CENTRAL) and the Cochrane COVID-19 study register.

Search strings for different searches are listed below.

Search of Cochrane Library - Issue 10 of 12, October 2020:

➢ #1 ((“step test” OR “walk test” OR “field test” OR “chair rise” OR “sit to stand” OR “exercise test” OR “exercise testing”)):ti,ab,kw OR (exertional NEAR/2 (desaturation OR hypoxia)):ti,ab,kw
➢ #2 MeSH descriptor: [Coronavirus Infections] this term only
➢ #3 (coronavirus OR covid*):ti,ab,kw
➢ #4 #2 or #3
➢ #5 #1 AND #4

Search of Cochrane COVID-19 study register:

➢ “step test” OR “walk test” OR “field test” OR “chair rise” OR “sit to stand” OR “exercise test” OR “exercise testing”

Our inclusion criteria used the PICOS/T frameworks as follows:

➢ *Population:* Individuals with Covid-19 or another lung disease with or without symptoms.
➢ *Interventions:* Any form of rapid exercise test performed at home or in a healthcare setting.
➢ *Comparator:* 6MWT or cardiopulmonary exercise test, CPET (both of which are used as gold standard in exercise testing), or a diagnostic test to diagnose the disease in question (eg bronchoalveolar lavage to diagnose pneumocystis carinii pneumonia).
➢ *Study* designs: clinical practice guidelines and systematic reviews addressing desaturation in exercise tests in lung disease, using the Cochrane definition of a Systematic Review. Primary human studies of all designs (e.g. experimental studies, quasi-experimental studies, diagnostic accuracy studies, and observational studies), excluding case series, that involved patients with COVID-19 or a lung disease undergoing rapid exercise testing were included. Editorials and letters around the subject of Covid-19 and rapid exercise testing were included to provide background and surface hypotheses on this new disease.
➢ *Outcome*. Pulse oximetry or arterial blood gas measurement, and association with any adverse outcome e.g. hospital admission, need for organ support, death.
➢ *Time periods:* All periods of time and duration of follow up.

No other limitations are imposed on the search or study selection process. Both peer-reviewed and preprint papers will potentially be eligible for inclusion. We plan to seek translation of any relevant papers published in languages other than English.

### Risk of bias appraisal

The risk of bias in individual studies will be assessed using the QUADAS 2 tool. Using the signalling questions, we will rate each potential source of bias as high, low, or unclear.

### Synthesis

The evidence synthetised from the search will be analysed through a formal statistical synthesis where possible. If the data is too heterogeneous for formal statistical analysis, it will be tabulated in summary tables of origins, methods and results, and then summarized narratively.

## Discussion

Covid-19 may cause an unusual form of acute lung disease characterised by a tendency to desaturate on minimal exertion. This rapid review aims to produce key findings relevant to the assessment of exertional desaturation in patients with suspected Covid-19. Establishing a validated tool to assess exertional desaturation will help to ensure that future research on this topic can be undertaken in a consistent way and that services can be planned to allow for appropriate planning of services to allow remote assessment (with the patient at home at a distance from the clinician) or assessment in an ambulatory setting such as a “hot hub” (where exercise tests may be performed outside in car parks, for example, for infection control reasons).

Risk to the patient from exercise tests should also be considered. Patients should be advised to terminate promptly if they develop any adverse symptoms (severe breathlessness, chest pain, dizziness) [23]. Tests involving climbing a flight of stairs should be avoided, since a staircase is a dangerous place to collapse.

## Data Availability

All data referred to in this manuscript is available.

## Declarations

### Ethics approval and consent to participate

Not applicable (desk research)

### Consent for publication

Not applicable (desk research)

### Availability of data and materials

All sources cited in this review are publicly available.

### Authors’ contributions

The conception of the work: TG, BJ, MK and MI-K. Designed the protocol and wrote the paper: AK, TG, BJ, MK and MI-K.

## Acknowledgements

We thank health informaticians and librarians: Helen F Williams, Jon Brassey and Nia Roberts.

